# Maternal dietary diversity and its correlates in a semi-urban municipality of Nepal: A cross-sectional study

**DOI:** 10.64898/2025.12.10.25342030

**Authors:** Reshika Rimal, Ayusha Rimal, Pranil Man Singh Pradhan

## Abstract

Maternal dietary diversity is vital for the health of both mothers and children during lactation, yet it is often compromised in low- and middle-income countries. This cross-sectional study among 251 lactating mothers in Tarakeswor Municipality, Nepal, assessed dietary diversity using a 24-hour dietary recall and the Minimum Dietary Diversity for Women (MDD-W) indicator. Overall, 68.1% of mothers achieved the minimum dietary diversity (≥5 of 10 food groups), with a mean score of 5.03 ± 1.25. In multivariable analysis, higher odds of meeting MDD were observed among mothers with secondary or higher education (aOR = 7.5; 95% CI: 3.8–15.0), employment (aOR = 2.9; 95% CI: 1.4–5.8), joint or extended family structure (aOR = 3.7; 95% CI: 1.9–7.0), the highest wealth quintile (aOR = 4.2; 95% CI: 1.9–9.1), food-secure households (aOR = 4.5; 95% CI: 2.3–7.9), adequate nutrition knowledge (aOR = 5.2; 95% CI: 2.7–9.8), ≥4 antenatal care visits (aOR = 1.9; 95% CI: 1.0–3.4), and higher empowerment (aOR = 3.9; 95% CI: 1.9–7.8). These findings highlight substantial socioeconomic disparities in maternal dietary diversity and underscore the need for integrated, equity-focused nutrition interventions in rapidly urbanizing settings in low- and middle-income countries.

## Introduction

Dietary diversity is defined as the number of different food groups consumed over a given reference period which is a cornerstone of adequate nutrition and a critical determinant of women’s health during the reproductive period [1–3]. It serves as a reliable proxy for overall diet quality and micronutrient adequacy, both of which are essential for sustaining maternal and child health [4, 5]. The Minimum Dietary Diversity for Women (MDD-W) indicator is defined as the consumption of at least five out of ten specified food groups in the previous 24 hours [6]. It is a validated measure used globally to assess the adequacy of women’s diets [7]. Ensuring that lactating mothers achieve this threshold is vital, as it directly influences the nutritional composition of breast milk, maternal recovery, and the growth and immune development of infants[8, 9].

However, in many low- and middle-income countries (LMICs), including Nepal, achieving adequate dietary diversity among reproductive women remains a persistent challenge [10, 11]. Maternal diets are often dominated by starchy staples such as rice, wheat, or maize, with limited inclusion of animal-source foods, legumes, fruits, and vegetables [12]. These patterns result from a complex interplay of economic constraints, limited food availability, and sociocultural norms that shape women’s access to and control over food resources [13, 14]. According to national survey data, more than half of Nepali women of reproductive age do not meet the MDD-W threshold, highlighting widespread dietary inadequacy and vulnerability to micronutrient deficiencies [15].

One of the primary barriers to dietary diversity is household food insecurity, which refers to limited or uncertain access to sufficient and nutritious food [16]. In food-insecure households, economic hardship often forces families to rely on inexpensive staple foods, leading to monotonous diets that lack micronutrient density [17]. Beyond economic limitations, cultural beliefs and postpartum dietary taboos also influence maternal food consumption [18]. For instance, a study in Bhaktapur, Nepal, documented that lactating mothers commonly avoided nutrient-rich foods, such as fish, green leafy vegetables, and eggs, due to the belief that these foods might adversely affect the infant [19]. These practices, though deeply rooted in local traditions, can exacerbate nutritional risks during a physiologically demanding period [20].

Environmental and structural challenges further restrict access to a diverse diet [21]. Seasonal food shortages, particularly during winter months, and increasing food prices limit the availability and affordability of perishable foods such as fruits, vegetables, and animal products [22]. Semi-urban and rural families are particularly affected, as they face both the economic pressures of urbanization and the resource limitations typical of rural settings [23]. This dynamic reflects a broader pattern across LMICs, where populations in transitional zones experience a “double burden” of undernutrition and emerging diet-related chronic diseases despite improvements in infrastructure and market access [24, 25].

Despite government and development efforts to strengthen maternal and child nutrition services in Nepal, undernutrition among women remains a major public health concern [26].The 2022 Nepal Demographic and Health Survey reported that 17% of women aged 15–49 years are underweight, with higher prevalence in rural and semi-urban areas [27]. While extensive research has examined maternal dietary patterns in rural and urban areas, semi-urban settings remain underexplored. These transitional environments are unique because they combine rural livelihoods and urban influences, creating a complex nutritional landscape shaped by rapid socioeconomic change, evolving food systems, and persistent inequality [28].

Understanding dietary diversity among lactating mothers in these contexts is vital for designing equitable nutrition programs. Semi-urban municipalities are home to a growing segment of Nepal’s population and represent a critical frontier for achieving Sustainable Development Goal 2, ending hunger and improving nutrition [29, 30]. Yet, most national policies and interventions continue to generalize urban and rural patterns, overlooking the nuanced realities of semi-urban families.

Therefore, this study aimed to assess the prevalence of minimum dietary diversity and identify its associated sociodemographic, economic, and household-level determinants, including food security, among lactating mothers in a semi-urban municipality of Nepal. By addressing this evidence gap, the study seeks to inform locally tailored, equity-driven interventions to promote maternal dietary diversity and improve health outcomes in rapidly urbanizing LMIC settings. To our knowledge, this is among the few studies focusing specifically on lactating women in Nepal’s transitional urban contexts, where rural and urban determinants of nutrition intersect.

## Methods

### Study Design and Setting

This was a community-based cross-sectional study conducted from September to November 2020 in Tarakeswor Municipality, a semi-urban area located in Kathmandu District, Nepal. Data collection and participant recruitment took place from 09/10/2020 to 24/03/2021. The municipality consists of 11 administrative wards and reflects both urban and rural characteristics, with a mix of socio-economic and cultural backgrounds [31]. According to municipal records, the estimated population of Tarakeswor was approximately 80,000 in 2020 [32].

### Study Population and Eligibility Criteria

The study population included lactating mothers with children under 24 months of age who were residing in the study area for at least six months. Eligible participants were women who were currently breastfeeding at the time of data collection. Mothers were excluded if they had a serious illness or were unable to respond to the interviewer-administered questionnaire due to physical or cognitive limitations.

A total of 700 eligible lactating mothers were identified across all 11 administrative wards of the Tarakeswor municipality, from which 251 mothers were selected using proportionate stratified random sampling [31]. Each ward served as a stratum, and the number of mothers selected was proportionate to the total number of lactating mothers identified in that ward. The ward-wise distribution and final sample allocation are detailed in the Sampling Method section.

### Sample Size and Sampling Method

The sample size was determined using the single population proportion formula [33]:

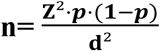

*Z* = 1.96 (standard normal value for 95% CI)

*p* = estimated prevalence of minimum dietary diversity (45%) from a similar study done in the periurban area of Nepal [19]

*d* = margin of error (5%)

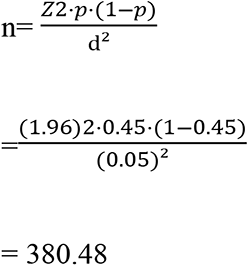

Since the total number of lactating mothers in the study area was below 10,000, the sample size was adjusted using the finite population correction formula:

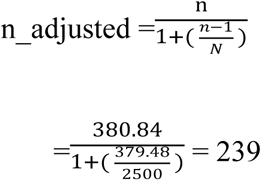

Finally, accounting for a 10% non-response rate:

Final N =239 + (239 * 0.1) = 263

A comprehensive sampling frame was created in coordination with Female Community Health Volunteers (FCHVs) and local health facilities, drawing from immunization records, growth monitoring charts, and household registers maintained by FCHVs to identify eligible participants. FCHVs are trained female community members who serve as primary health liaisons between the government health system and local populations, playing a critical role in maternal and child health service delivery in Nepal [34] . The number of lactating mothers was determined in each ward, and the sample was proportionately allocated. Within each ward, participants were selected using simple random sampling to ensure equal probability of selection and minimize selection bias. The ward-wise distribution and final sample allocation are shown in **Table 1**.

**Table 1:** Ward-wise Distribution of Total Lactating Mothers and Sample Size Selected (n=263).

| Ward Number | Total Lactating Mothers | Sample Size Selected |
| --- | --- | --- |
| Ward 1 | 55 | 21 |
| Ward 2 | 60 | 23 |
| Ward 3 | 40 | 15 |
| Ward 4 | 75 | 28 |
| Ward 5 | 30 | 11 |
| Ward 6 | 120 | 45 |
| Ward 7 | 90 | 34 |
| Ward 8 | 45 | 17 |
| Ward 9 | 65 | 24 |
| Ward 10 | 50 | 19 |
| Ward 11 | 70 | 26 |
| **Total** | **700** | **263** |

Although 263 mothers were selected, only 251 completed the interviews, resulting in a response rate of 95.4%. Twelve mothers either declined to participate or were unavailable despite repeated follow-up visits. These individuals were excluded from the final analysis.

## Study Variables

### Outcome Variable

#### Dietary Diversity

Dietary diversity was assessed using the Minimum Dietary Diversity for Women (MDD-W) indicator developed by the FAO [6]. A 24-hour dietary recall method was used to record all foods and beverages consumed by participants. Food items were categorized into ten predefined food groups: starchy staples, dark green leafy vegetables, vitamin A-rich fruits and vegetables, other vegetables, other fruits, dairy, meat, poultry, fish, eggs, legumes, nuts, seeds, oils, and fats. Women who consumed foods from five or more of these food groups were considered to have achieved minimum dietary diversity, while those who consumed fewer than five groups were categorized as not achieving it [35] .

### Independent variables: Sociodemographic Variables

These included maternal age (in completed years), educational status (no formal education, primary, secondary, or higher), and employment status (employed or unemployed). Household type was classified as either joint or nuclear, and monthly household income was categorized into tertiles (low, middle, and high). These classifications were based on standard frameworks used in the Nepal Demographic and Health Survey [27].

#### Wealth Quintile

Household wealth status was assessed using a composite wealth index, constructed through Principal Component Analysis (PCA). Variables included household assets (e.g., radio, mobile phone, refrigerator), housing structure (floor and roof materials), type of cooking fuel, and access to electricity, drinking water, and sanitation. Based on PCA scores, households were categorized into five quintiles: poorest, poorer, middle, richer, and richest. This method followed the Nepal Demographic and Health Survey [27].

#### Nutrition Knowledge

Maternal knowledge of nutrition during lactation was assessed using ten structured questions, adapted from the Food and Agriculture Organization (FAO) Knowledge, Attitudes, and Practices (KAP) framework[36] . Questions covered topics such as dietary diversity, sources of essential nutrients, iron-folic acid supplementation, and the consequences of undernutrition. Each correct response received one point. Participants scoring ≥5 out of 10 were categorized as having adequate knowledge, while those scoring <5 were categorized as having inadequate knowledge. This classification was based on standard frameworks used in the Nepal Demographic and Health Survey [27].

#### Household Food Insecurity Access Scale (HFIAS)

(HFIAS), a standardized tool developed by the Food and Nutrition Technical Assistance (FANTA) Project [37]. It includes nine structured questions that evaluate experiences related to uncertainty about food supply, insufficient food quality, and inadequate food intake over the previous 30 days. Each question was followed by a frequency-of-occurrence item scored as 0 (never), 1 (rarely), 2 (sometimes), or 3 (often), resulting in a total score ranging from 0 to 27, with higher scores indicating more severe food insecurity. Based on the total score, households were initially categorized into four groups: food secure, mildly food insecure, moderately food insecure, and severely food insecure. For analytical purposes, this variable was further dichotomized into two categories: food secure (including only food secure) and food insecure (combining mildly, moderately, and severely food insecure households). This method followed the Nepal Demographic and Health Survey [27].

#### Women’s Empowerment

Women’s empowerment was assessed using key decision-making indicators adapted from the Nepal Demographic and Health Survey (NDHS) [27]. Respondents were asked whether they participated in decision-making regarding their own health care, major household purchases, and visits to family or relatives. Responses were scored and summed, and participants were categorized into two groups: empowered (participated in one or more decisions) and not empowered (did not participate in any of the decisions). This variable was used to assess the role of autonomy and agency in influencing maternal dietary diversity.

#### Study Procedure

Data for this study were collected using a semi-structured interviewer-administered questionnaire that was developed by the researcher based on previously validated tools and guidelines. The questionnaire was initially prepared in English and translated into Nepali to ensure clarity, cultural appropriateness, and ease of understanding for the participants. The full questionnaire is available in the Supplementary File. It was pretested among a similar group of lactating mothers in a neighboring municipality, and minor modifications were made based on the feedback received to improve the clarity and flow of the questions.

The data collectors were two trained medical students. They received detailed training on standardized interviewing techniques and ethical research practices for 2 days. Enumerator training also included role play and field simulation to ensure consistency in data collection. The final version of the questionnaire was piloted, refined, and used with 15 lactating mothers in a neighboring ward. Necessary modifications were made to improve clarity.

#### Data Analysis

Data were entered into EpiData and analyzed using SPSS version 26. Descriptive statistics, including means, standard deviations, and proportions, were used to summarize participant characteristics. Bivariate logistic regression was conducted to assess the association between independent variables and dietary diversity. Variables with p-values < 0.20 in the bivariate analysis were entered into the multivariable logistic regression model. Adjusted odds ratios (aORs) with 95% confidence intervals (CIs) were reported. All variables included in the final model were tested for multicollinearity, and none were found to exceed the threshold (VIF < 2). Model fit was assessed using the Hosmer–Lemeshow goodness-of-fit test and was acceptable (p = 0.48), indicating a good fit to the observed data. Statistical significance was set at p < 0.05

## Results

### Participant Characteristics

Table 2 summarizes the demographic, household, and health-related characteristics of the 251 lactating mothers included in the study. Among the 251 lactating mothers included in the study, most were aged 18–25 years (69.7%), with smaller proportions aged 26–35 years (28.3%) and above 35 years (2.0%). More than half of the respondents belonged to upper-caste groups (54.6%), followed by disadvantaged Janajati (26.7%) and Dalit communities (17.1%). The majority of participants identified as Hindu (95.6%). Three-fourths of the mothers lived in joint or extended family settings (75.3%), and most households were male-headed (68.5%). Land ownership was common, with 70.9% of households owning land.

**Table 2:**
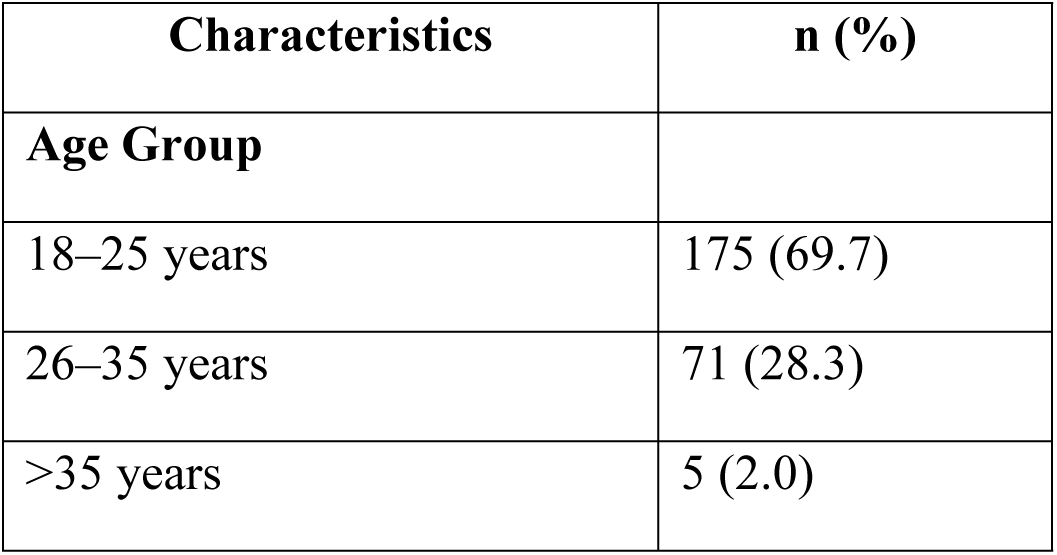
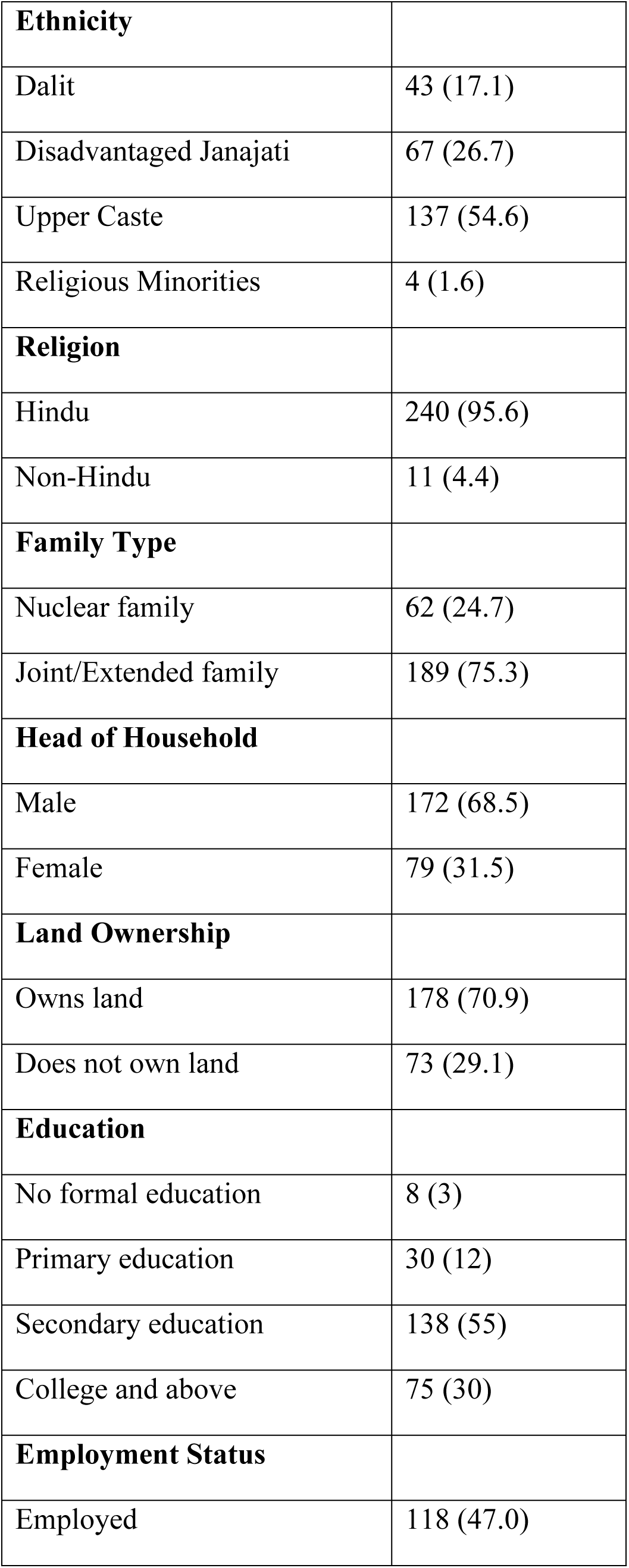
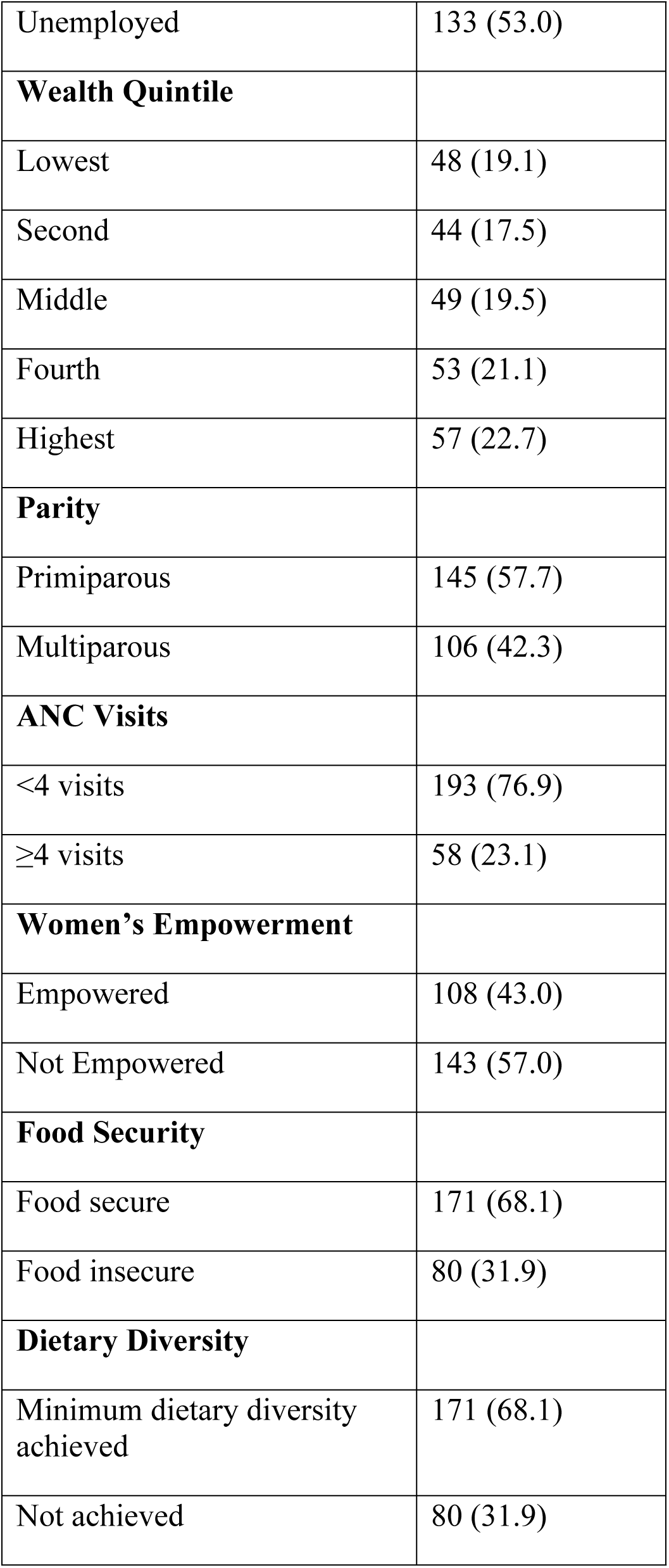
Key Characteristics of the Study Population (n = 251)

Regarding education, 3% of mothers had no formal schooling, 12% had completed primary education, over half (55%) had secondary education, and 30% had attained college-level or higher education. Nearly half of the women were employed (47%). Wealth distribution was relatively even across quintiles, with 22.7% in the highest and 19.1% in the lowest quintile. More than half of the mothers were primiparous (57.7%).

In terms of healthcare utilization, only 23.1% had completed four or more antenatal care (ANC) visits. Regarding empowerment, 43% of women were categorized as empowered. Household food security was reported by 68.1% of participants. A similar proportion of mothers (68.1%) achieved minimum dietary diversity, with a mean score of 5.03 (SD ± 1.25).

### Dietary Diversity Among Lactating Mothers

The primary outcome of interest was dietary diversity, assessed using the Minimum Dietary Diversity for Women (MDD-W) indicator. Among the 251 lactating mothers, 68.1% (n = 171) met the minimum dietary diversity threshold by consuming foods from at least five out of ten defined food groups in the previous 24 hours. The mean dietary diversity score was 5.03 (SD ± 1.25). However, 31.9% (n = 80) of participants did not achieve minimum dietary diversity, indicating potential nutritional inadequacies in nearly one-third of the population.

To further explore the types of foods consumed, **Table 3** presents the proportion of lactating mothers who consumed each of the ten food groups within the past 24 hours based on the MDD-W framework. All participants consumed starchy staples such as rice, maize, and potatoes, which are culturally ingrained and widely accessible. Similarly, pulses were consumed by nearly all mothers (98.8%), reflecting their affordability and routine inclusion in traditional Nepali meals.

**Table 3:**
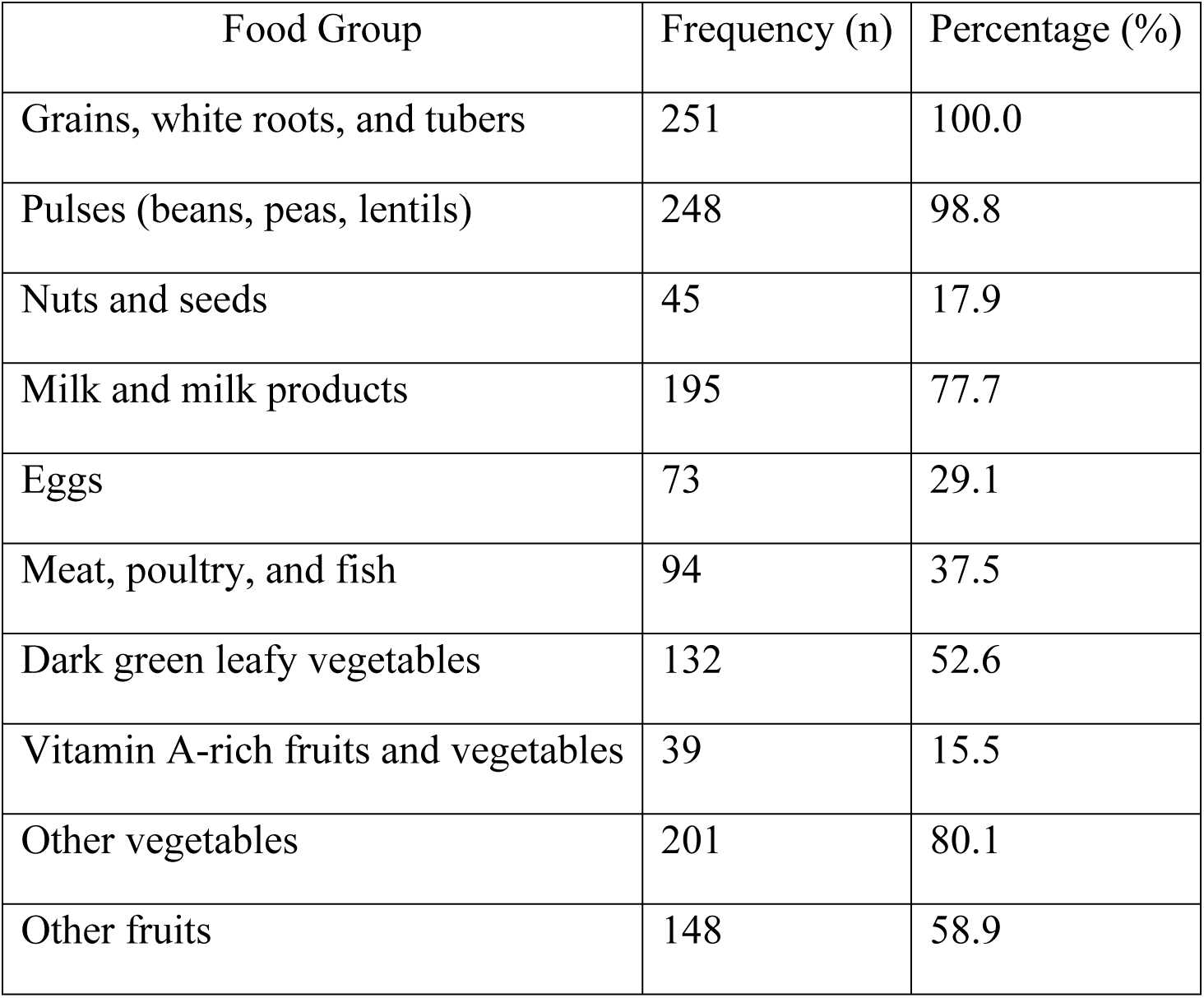
Proportion of lactating mothers who consumed each of the 10 food groups within the past 24 hours, based on the MDD-W framework (N=251).

| Food Group | Frequency (n) | Percentage (%) |
| --- | --- | --- |
| Grains, white roots, and tubers | 251 | 100.0 |
| Pulses (beans, peas, lentils) | 248 | 98.8 |
| Nuts and seeds | 45 | 17.9 |
| Milk and milk products | 195 | 77.7 |
| Eggs | 73 | 29.1 |
| Meat, poultry, and fish | 94 | 37.5 |
| Dark green leafy vegetables | 132 | 52.6 |
| Vitamin A-rich fruits and vegetables | 39 | 15.5 |
| Other vegetables | 201 | 80.1 |
| Other fruits | 148 | 58.9 |

In contrast, the intake of several nutrient-dense food groups was substantially lower. Only 29.1% of mothers consumed eggs, 17.9% consumed nuts and seeds, and just 15.5% consumed vitamin A-rich fruits and vegetables. These items, although vital sources of high-quality protein, essential fatty acids, and micronutrients such as vitamin A and iron, may be less consumed due to multiple factors. These include economic constraints, cultural taboos during the postpartum period, seasonal unavailability, and limited nutrition awareness. Despite most mothers meeting the minimum diversity threshold, these findings highlight the risk of “hidden hunger” and suggest a need for targeted nutrition interventions to promote affordable, locally available, and micronutrient-rich foods in the diets of lactating women.

### Factors Associated with Minimum Dietary Diversity Among Lactating Mothers

Table 4 presents the multivariable logistic regression results for factors associated with achieving minimum dietary diversity among lactating mothers in Tarakeswor Municipality, Nepal. In the adjusted model, several sociodemographic and maternal characteristics remained significant predictors.

**Table 4.** Multivariable Logistic Regression of Factors Associated with Minimum Dietary Diversity Among Lactating Mothers in Tarakeswor Municipality, Nepal (n = 251)

| Variables | Not Diverse<br>n (%) | Diverse n<br>(%) | Crude Odds Ratio<br>(95% CI) | Adjusted Odds<br>Ratio (95% CI) |
| --- | --- | --- | --- | --- |
| **Age Groups** |  |  |  |  |
| ≤24 years | 90 (36.0) | 161 (64.0) | Ref | Ref |
| >24 years | 40 (47.5) | 44 (52.5) | 1.60 (0.95–2.65) | 1.30 (0.70–2.20) |
| **Ethnicity** |  |  |  |  |
| Dalit | 26 (57.0) | 20 (43.0) | Ref | Ref |
| Janajati | 35 (37.0) | 60 (63.0) | 2.10 (1.10–4.00)* | 1.80 (0.90–3.50) |
| Brahmin/Chhetri | 32 (25.0) | 96 (75.0) | 3.80 (1.90–7.20)** | 3.10 (1.50–6.50)* |
| **Family Type** |  |  |  |  |
| Nuclear | 72 (50.0) | 72 (50.0) | Ref | Ref |
| Joint/Extended | 49 (21.0) | 184 (79.0) | 4.50 (2.70–7.70)** | 3.70 (1.90–7.00)* |
| **Education Level** |  |  |  |  |
| Secondary or Lower | 43 (72.0) | 17 (28.0) | Ref | Ref |
| SLC or Above | 78 (30.0) | 182 (70.0) | 9.00 (4.60–17.50)** | 7.50 (3.80–15.00)<br>** |
| **Employment** |  |  |  |  |
| Unemployed | 97 (46.0) | 114 (54.0) | Ref | Ref |
| Employed | 24 (20.0) | 96 (80.0) | 3.40 (1.80–6.60) * | 2.90 (1.40–5.80) * |
| **Wealth Status** |  |  |  |  |
| Lowest | 42 (56.0) | 33 (44.0) | Ref | Ref |
| Middle | 38 (39.0) | 58 (61.0) | 2.10 (1.00–4.20) | 1.80 (0.90–3.60) |
| Highest | 13 (19.0) | 55 (81.0) | 5.00 (2.30–10.60) ** | 4.20 (1.90–9.10) * |
| **Food Security** |  |  |  |  |
| Food Insecure | 48 (65.0) | 26 (35.0) | Ref | Ref |
| Food Secure | 73 (26.0) | 207 (74.0) | 5.10 (2.80–8.90)** | 4.50 (2.30–7.90)** |
| **Nutrition Knowledge** |  |  |  |  |
| Inadequate | 62 (58.0) | 45 (42.0) | Ref | Ref |
| Adequate | 59 (23.0) | 198 (77.0) | 6.00 (3.10–11.00)** | 5.20 (2.70–9.80)** |
| **ANC Visits** |  |  |  |  |
| <4 | 75 (52.2) | 27 (47.8) | Ref | Ref |
| ≥4 | 28 (34.2) | 71 (65.8) | 2.00 (1.10–3.60) * | 1.90 (1.00–3.40)* |
| **Women**<br>**Empowerment** |  |  |  |  |
| Not Empowered | 64 (44.8) | 79 (55.2) | Ref | Ref |
| Empowered | 16 (14.8) | 92 (85.2) | 4.67 (2.51–8.67) ** | 3.90 (1.90–7.80)** |

cOR = Crude Odds Ratio; aOR = Adjusted Odds Ratio; Ref = Reference category.
\*Significance levels: \*p < 0.05, \*\*p < 0.001

Mothers residing in joint or extended families had significantly higher odds of meeting the minimum dietary diversity compared to those living in nuclear families (aOR = 3.7; 95% CI: 1.9–7.0). Similarly, those with secondary or higher education were substantially more likely to achieve dietary diversity than mothers with no formal or only primary education (aOR = 7.5; 95% CI: 3.8–15.0). Employment status was also an important determinant - employed women had almost three times higher odds of achieving minimum dietary diversity compared to unemployed women (aOR = 2.9; 95% CI: 1.4–5.8).

Household socioeconomic position showed a clear gradient effect. Women from the highest wealth quintile were over four times more likely to achieve adequate dietary diversity than those in the lowest quintile (aOR = 4.2; 95% CI: 1.9–9.1). Similarly, food security was strongly associated; mothers from food-secure households had 4.5-fold greater odds of achieving dietary diversity compared with those from food-insecure households (aOR = 4.5; 95% CI: 2.3–7.9).

Maternal nutrition knowledge emerged as another key factor. Women with adequate knowledge had over five times higher odds of meeting dietary diversity than those with inadequate knowledge (aOR = 5.2; 95% CI: 2.7–9.8). Moreover, attending four or more antenatal care (ANC) visits was associated with better dietary diversity (aOR = 1.9; 95% CI: 1.0–3.4). Finally, women’s empowerment significantly increased the likelihood of achieving dietary diversity compared to those who were not empowered (aOR = 3.9; 95% CI: 1.9–7.8).

## Discussion

This study provides important evidence on maternal dietary diversity in a transitional urban setting in Nepal, where rural and urban dynamics coexist and interact. As low- and middle-income countries (LMICs) experience rapid urbanization, understanding maternal nutrition in these hybrid environments is essential for designing equitable public health interventions [38]. Tarakeswor Municipality represents such a transitional zone, exhibiting infrastructural growth while still grappling with socioeconomic disparities [32]. Our findings thus offer insights not only for Nepal but also for other LMICs facing similar urban transformations. This cross-sectional study found that a substantial proportion of lactating mothers in Tarakeswor Municipality did not meet the minimum dietary diversity recommended by the MDD-W framework. Specifically, 68.1% of mothers achieved minimum dietary diversity (≥5 food groups), while 31.9% did not, indicating the presence of potential nutritional inadequacies in nearly one-third of the population.

Dietary diversity among lactating mothers in South Asia remains suboptimal despite its critical importance for maternal and child health [39, 40, 40, 41]. Evidence from Nepal illustrates this concern: in remote areas, only about 28% of lactating women meet the minimum dietary diversity (MDD-W), while even in the more developed Kathmandu Valley, less than 45% reach this threshold [19, 42]. These figures reflect broader trends observed across semi-urban South Asia, where maternal diet quality is shaped by complex interactions among socioeconomic, cultural, and behavioral factors [25, 43, 44].

In our study of lactating mothers in Tarakeswor Municipality, the mean Dietary Diversity Score (DDS) was 5.03 (±1.25), with 68% of mothers meeting the MDD-W threshold. This is higher than what has been reported in some rural Nepali settings, such as Bajhang (DDS = 4.1) [42], but lower than in neighboring countries like Bangladesh and Vietnam , where robust maternal nutrition programs exist [41, 45]. Furthermore, structural barriers such as urban poverty and limited access to healthy diets have been identified as persistent challenges to achieving dietary adequacy in LMICs [23] . This underscores the semi-urban paradox: while access to food markets and health services may be better than in remote regions, structural barriers such as poverty, low education, and gender norms still restrict optimal dietary practices [46, 47].

Our findings confirm several key predictors of dietary diversity. Maternal education emerged as a powerful determinant: women with secondary or higher education were 7.5 times more likely to meet the MDD-W threshold compared to those with no or primary education. This is consistent with national survey data (NDHS, 2022) [27]and studies from across South Asia and Africa [39, 48, 49] . In a study conducted in Baglung Municipality, women with higher education had significantly greater dietary diversity, reinforcing the notion that education enhances awareness, decision-making, and access to a varied diet [10].

Similarly, employment status was positively associated with dietary diversity. Employed mothers were nearly three times more likely to achieve the MDD-W threshold. Employment provides not just income but also autonomy, enabling women to make independent food choices and purchase nutrient-dense foods. In a study conducted in Nepal, employed women had nearly five times the odds of achieving dietary diversity [10]. These findings highlight the need to promote economic opportunities for women as part of nutrition interventions.

Household wealth and food security were also critical. Mothers from the highest wealth quintile had more than four times higher odds of achieving dietary diversity than those from the lowest quintile, supporting Bennett’s Law that income growth leads to more diverse diets [50]. Food-secure households were almost four times more likely to meet the MDD-W threshold, aligning with findings from Ethiopia, Bangladesh, and Nepal [39, 42, 51]. In Baglung Municipality, Nepal, women in the highest wealth group had more than three times the odds of achieving dietary diversity, and food insecurity was closely associated with monotonous diets [10].

Family structure also influenced dietary outcomes. Women living in joint or extended families were significantly more likely to achieve adequate dietary diversity (AOR = 3.7), possibly due to shared food resources and household support systems. While some African studies show the opposite effect due to intra-household food allocation challenges in larger families where limited resources must be divided among more members, South Asian contexts often demonstrate the benefits of communal support [10, 52–54]. In these settings, extended family structures may buffer food insecurity by pooling income, sharing meals, and providing social or caregiving support to women during lactation and child-rearing.

Nutrition knowledge was one of the strongest predictors of dietary diversity in our study (AOR = 5.2). Women who understood the importance of varied diets during lactation, sources of essential nutrients, and the consequences of undernutrition were more likely to make healthier food choices. This finding aligns with global evidence showing that education and counseling interventions significantly improve dietary outcomes[55, 56].

Lastly, health service utilization, particularly antenatal care (ANC), was positively associated with dietary diversity. Women who had four or more ANC visits had nearly twice the odds of achieving dietary diversity, although the association did not reach strong statistical significance. This finding is consistent with literature from Ethiopia and South Asia showing that ANC visits provide critical opportunities for nutrition education and behavior change [41, 42, 51, 57].

While ethnicity was significant in the bivariate analysis, it was not retained in the multivariable model. This may reflect that, in semi-urban settings like Tarakeswor, educational and economic factors outweigh ethnic disparities that are more pronounced in rural regions. A similar study in Nepal observed that socioeconomic and empowerment factors had stronger predictive value than ethnicity [10, 19].

Overall, our findings align with regional research, demonstrating the convergence of evidence around key determinants of dietary diversity, namely education, employment, wealth, household structure, food security, nutrition knowledge, and health service utilization. The consistency of these findings across studies suggests that multi-sectoral strategies addressing these factors can effectively enhance maternal nutrition in Nepal and similar LMIC contexts. These findings support national nutrition policies such as Nepal’s Multi-Sector Nutrition Plan and can inform targeted interventions in comparable settings undergoing rapid urban transitions [26].

## Conclusion

This study showed that although 68.1% of lactating mothers in Tarakeswor Municipality met the minimum dietary diversity (MDD-W ≥ 5), nearly one-third did not, highlighting persistent nutritional vulnerabilities even in areas with access to urban services. Food insecurity was the strongest barrier to adequate diet quality, further compounded by low maternal education, unemployment, limited household wealth, inadequate nutrition knowledge, and restricted decision-making autonomy. As one of the few studies focusing on lactating mothers in a semi-urban, transitional municipality of Nepal, this research adds new evidence that urban proximity alone does not ensure dietary adequacy; social and economic empowerment remain central to improving maternal nutrition.

Public health efforts should therefore integrate equity-focused and multi-sectoral interventions that strengthen women’s education, economic participation, and household food security, alongside routine nutrition counseling during antenatal and postpartum care. Addressing these interconnected factors is essential for advancing Nepal’s maternal health and nutrition goals and achieving Sustainable Development Goals 2 (Zero Hunger) and 3 (Good Health and Well-Being).

## Strengths and Limitations

This study is among the first to examine maternal dietary diversity specifically among lactating mothers in a semi-urban, transitional municipality of Nepal, where rural and urban dynamics intersect. Most previous research in Nepal has focused either on rural or metropolitan settings; by contrast, this study contributes new evidence from an understudied population experiencing rapid socioeconomic transition.

A key strength lies in its community-based design using proportionate stratified random sampling across all municipal wards, ensuring representativeness and minimizing selection bias. The study also utilized validated international tools the Minimum Dietary Diversity for Women (MDD-W) and Household Food Insecurity Access Scale (HFIAS)—allowing comparability with global and national datasets. Moreover, by incorporating variables such as nutrition knowledge, empowerment, and antenatal care utilization, it provides a more holistic understanding of the social determinants of maternal diet quality in Nepal.

However, several limitations should be noted. The cross-sectional design precludes causal inference. The use of a single 24-hour dietary recall may not capture habitual intake and is subject to recall bias. Measures of food security, empowerment, and knowledge were self-reported, which may introduce social desirability bias. Additionally, because the study was conducted in a single semi-urban municipality, the findings may not be generalizable to other ecological or cultural settings in Nepal. Seasonal variation and cultural dietary restrictions were not systematically captured, and the operationalization of empowerment as participation in household decision-making may not fully reflect its multidimensional nature.

## Abbreviations

ANC: Antenatal Care
aOR: Adjusted Odds Ratio
CI: Confidence Interval
DDS: Dietary Diversity Score
FCHV: Female Community Health Volunteer
FAO: Food and Agriculture Organization
HFIAS: Household Food Insecurity Access Scale
LMICs: Low- and Middle-Income Countries
MDD-W: Minimum Dietary Diversity for Women
NDHS: Nepal Demographic and Health Survey
PCA: Principal Component Analysis
SPSS: Statistical Package for the Social Sciences
WHO: World Health Organization

## Ethics approval and consent to participate

Ethical approval for the study was obtained from the Institutional Review Committee (IRC) of the Institute of Medicine, Tribhuvan University, Nepal (Approval No. 86(6-1) E2/077/078).

Written informed consent was obtained from all participants. For participants below the age of 20, written assent was obtained from the participant, and consent was obtained from a parent or guardian, following IRC ethical protocols. Anonymity and confidentiality were ensured throughout the study. This study was conducted in accordance with the ethical principles outlined in the Declaration of Helsinki.

## Consent for publication

Not applicable.

## Availability of data and materials

All relevant data supporting the findings of this study are provided within the manuscript and its Supporting Information files.

## Acknowledgments

We gratefully acknowledge the data collectors for their support during fieldwork and the Central Department of Public Health, Tribhuvan University, for their institutional guidance. We also thank the lactating mothers who participated in the study and the Female Community Health Volunteers (FCHVs) for their assistance in the community engagement process.

